# Evaluation of automated pediatric sleep stage classification using U-Sleep - a convolutional neural network

**DOI:** 10.1101/2024.08.18.24312174

**Authors:** Ajay Kevat, Rylan Steinkey, Sadasivam Suresh, Warren R Ruehland, Jasneek Chawla, Philip I Terrill, Andrew Collaro, Kartik Iyer

## Abstract

**Study Objectives:** U-Sleep is a publicly-available automated sleep stager, but has not been independently validated using pediatric data. We aimed to a) test the hypothesis that U-Sleep performance is equivalent to trained humans, using a concordance dataset of 50 pediatric polysomnogram excerpts scored by multiple trained scorers, and b) identify clinical and demographic characteristics that impact U-Sleep accuracy, using a clinical dataset of 3114 polysomnograms from a tertiary center.

**Methods:** Agreement between U-Sleep and ‘gold’ 30-second epoch sleep staging was determined across both datasets. Utilizing the concordance dataset, the hypothesis of equivalence between human scorers and U-Sleep was tested using a Wilcoxon two one-sided test (TOST). Multivariable regression and generalized additive modelling were used on the clinical dataset to estimate the effects of age, comorbidities and polysomnographic findings on U-Sleep performance.

**Results:** The median (interquartile range) Cohen’s kappa agreement of U-Sleep and individual trained humans relative to “gold” scoring for 5-stage sleep staging in the concordance dataset were similar, kappa=0.79(0.19) vs 0.78(0.13) respectively, and satisfied statistical equivalence (TOST p<0.01). Median (interquartile range) kappa agreement between U-Sleep 2.0 and clinical sleep-staging was kappa=0.69(0.22). Modelling indicated lower performance for children <2 years, those with medical comorbidities possibly altering sleep electroencephalography (kappa reduction=0.07-0.15) and those with decreased sleep efficiency or sleep-disordered breathing (kappa reduction=0.1).

**Conclusion:** While U-Sleep algorithms showed statistically equivalent performance to trained scorers, accuracy was lower in children <2 years and those with sleep-disordered breathing or comorbidities affecting electroencephalography. U-Sleep is suitable for pediatric clinical utilization provided automated staging is followed by expert clinician review.

## Introduction

Sleep in children is important for growth and learning,^1,2^ and disorders that disrupt sleep during childhood may have significant consequences on neurobehavioral and somatic growth functions. Specifically, impaired sleep in childhood is associated with cognitive dysfunction,^3^ emotional dysregulation,^4,5^ behavioral problems,^6,7^ and metabolic syndromes.^8,9,10^ Early identification of conditions that disrupt sleep in children enables prompt treatment that has been shown to remediate behavioral, cognitive and metabolic dysfunction.^11,12^ Diagnostic polysomnography (PSG), involving recording of cardiorespiratory and neurophysiologic signals during sleep, is the gold standard test for identifying most sleep disorders in children.^13,14^ Review of PSG data is a laborious process performed by a trained pediatric sleep scientist and/or physician. Sleep staging is an important component of the review process, which can impact upon the diagnostic conclusions made by the PSG that guide subsequent clinical management. Unfortunately, sleep staging is both time-consuming and subject to significant human inter-observer variation,^15^ despite well-defined scoring rules developed to standardize scoring between clinicians and centers.^16^

To address the demands on clinician time and the subjectivity of sleep staging in PSG, machine learning algorithms to automate sleep staging have been developed. These include algorithms developed specifically for children of varying ages,^17–25^ due to differences in brainwave morphology at different stages of development,^26^ and because adult sleep staging algorithms are known to be less performant when applied to children.^27^ However, these pediatric-specific sleep staging algorithms are not freely available. U-Sleep provides state-of-the-art, sleep staging algorithms that offer an automated approach to pediatric sleep staging, by leveraging a fully convolutional neural network which is trained on a combination of large adult ± pediatric datasets, and is freely available for scientific and non-commercial use.^28^ However, to date, U-Sleep’s performance when applied to unseen pediatric datasets has yet to be independently tested and evaluated.

The overarching aim of this study was to independently test publicly available versions of U-Sleep and compare the performance of these models to clinician expert scoring, to determine if the algorithm(s) would be suitable for clinical use in pediatric populations. We address this aim with two specific objectives:

1. *Compare the performance of the U-Sleep algorithm to individual human inter-scorer performance, using a trained expert majority score as the gold standard.* This objective is addressed using a “concordance dataset” of 50 pediatric PSG excerpts each scored by multiple (median 20) trained and in-training human scorers. We hypothesized that U-Sleep and trained clinicians would perform equivalently when compared to sleep staging determined by expert consensus.
2. *Identify key clinical or demographic characteristics that impact the accuracy of U-Sleep.* This objective is addressed using a “clinical dataset” of 3114 diagnostic PSGs from a single tertiary hospital. U-Sleep sleep staging is compared to the clinical sleep staging performed by pediatric sleep staff, with multivariable regression analysis conducted to assess the influence of demographic and clinical characteristics (age, sleep disordered breathing severity, clinical diagnosis associated with abnormal electroencephalography (EEG)) on U-Sleep performance.

## Methods

We assessed the performance of U-Sleep algorithms for pediatric sleep staging using two datasets. Both were scored using methodology outlined by the American Academy of Sleep Medicine.^16^

### Concordance Dataset

The concordance dataset included 50 pediatric PSG excerpts (each 95 minutes in length) with a total of 9516 (553 Wake, 215 N1, 4080 N2, 2510 N3 and 2158 REM) 30-second epochs used for Australian and international concordance exercises, operated by QSleep (qsleep.com.au). Sleep staging of each PSG excerpt (from a pediatric patient, with specific age unknown and no identifiable information contained within) was performed by varying numbers of human scorers from different pediatric sleep centers, with the majority being from Australia. Most scorers were clinicians with substantial experience in pediatric sleep staging (trained scorers), and a minority were human scorers from the same centers classified as in-training scorers. We extracted the number of trained and in-training human scorers who contributed to each of the PSG excerpts and summarized these data. The ‘gold’ sleep stage for each 30-second epoch was the most frequently selected stage (i.e. mode) by trained scorers. For each study, electrooculogram (EOG) and EEG channels were extracted from the 50 PSG excerpts in European Data Format (EDF) and the corresponding signals were staged in 30-second epochs by each of three available U-Sleep algorithms (U-Sleep 1.0 (EEG + EOG), U-Sleep 2.0 (EEG + EOG), and U-Sleep 2.0 EEG) via sequential upload at https://sleep.ai.ku.dk. We assessed comparative performance of each of the three EEG leads (frontal, central, occipital) by uploading them separately. We assessed agreement between the U-Sleep algorithm outputs and the ‘gold’ sleep stages for all available epochs in the dataset by calculating percentage simple agreement and Cohen’s kappa when utilizing a five-stage approach (Wake, Rapid Eye Movement (REM), N1, N2, N3), three-stage approach (Wake, REM, non-REM) and two-stage approach (Wake/Sleep). We similarly assessed agreement achieved by trained, in-training, and pooled (i.e. both trained and in-training) human scorers. To address our primary hypothesis that U-Sleep algorithm performance for clinical sleep staging is equivalent to expert clinicians, we tested for equivalence using a Wilcoxon two one-sided test (TOST), with trained scorer interquartile range (IQR) used as the equivalence bound. We also determined correct and incorrect sleep stage classification rates for each sleep stage by each U-Sleep algorithm and converted these into confusion matrices. For statistical analysis of the concordance dataset, Python v3.10.14 (with pandas v2.1.4 and scikit-learn v1.3.0) was used; seaborn v0.12.2 and matplotlib v3.8.4 were utilized to generate figures.

### Clinical Dataset

The second dataset consisted of 3114 diagnostic PSG studies (with approximately 3.3 million epochs) performed at a single tertiary pediatric center (clinical dataset; see Table S1 in Supplemental Material). Sleep staging of each epoch was performed by a pediatric sleep nurse overnight during the night of data capture and this was reviewed and revised by a pediatric sleep physician; the final revised sleep stage assigned to each epoch was considered the ‘gold’ sleep stage for this dataset. Children <3 months of age at the time of PSG were excluded given staging is usually performed using active and quiet sleep staging in this group.^29^ When staging the clinical dataset, U-Sleep 2.0 was used, given this has been trained on more pediatric data than its predecessor. To enable batch processing of the entire clinical dataset, we used the U-Sleep Webserver API and associated Python bindings (https://github.com/perslev/U-Sleep-API-Python-Bindings). We packaged each individual data file as de-identified temporary EDF files and, via the U-Sleep Webserver, permitted the algorithm to consider all combinations of EEG input channel configurations; the algorithm is designed so that each combination will be used for prediction one at a time, and then a majority voting is computed across all predictions to produce the final sleep staging. Once scored by the algorithm, the U-Sleep scored file was exported and saved and the temporary EDF file deleted, with U-Sleep scored labels used for comparison with ‘gold’ labels. This process was completed for all 3114 files. We calculated agreement (Cohen’s kappa) between the ‘gold’ labels and U-Sleep when utilizing a five-, three- and two-stage approach.

As the clinical dataset contained demographic, PSG, and clinical information for most patients, we analyzed how agreement (Cohen’s kappa) varied with patient age, primary diagnosis grouped into categories, and PSG findings related to sleep disordered breathing (SDB) diagnosis and sleep efficiency, using a combination of ordinary least squares (OLS) regression and generalized additive modelling. Cohen’s kappa was modelled separately with each factor using OLS univariable modelling, and together using an OLS multivariable model. Primary diagnosis was categorized into three; Group 1 (n = 13) - diagnosis highly likely to manifest with sleep EEG abnormalities (i.e. Rett syndrome, epileptic encephalopathy), Group 2 (n = 242) - diagnosis that may cause sleep EEG abnormalities (i.e. complex chromosomal or genetic disorder such as Trisomy 21), and Group 3 (n = 2539) - remaining children (inclusive of those with autism spectrum disorder). SDB diagnostic categories were central (central apnea hypopnea index (CAHI) ≥5; n = 200), moderate-severe obstructive (obstructive apnea hypopnea index (OAHI) ≥5, n = 394), mixed (CAHI ≥5 and OAHI ≥5, n = 81), and none (n = 2119); each participant belonged to only one of these categories. Sleep efficiency was defined as percentage of PSG analysis time spent asleep. R (R Core Foundation) 4.2.3 and MATLAB 2022a (The MathWorks Inc, Natick, MA) was used for analyses and data visualization, including estimation of Cohen’s kappa and calculation of simple agreement (percent agreement of correct classifications divided by the total number of observations).

## Results

### Concordance dataset

The median number of trained scorers who participated in the concordance exercise for each of the 50 PSG excerpts (and therefore contributed to the formation of the ‘gold’) was 16 (IQR 5, range 6-24). The median number of in-training scorers who partook in the concordance exercise for each of the PSG excerpts was four (IQR 3.5, range 1-13). Table 1 and Table S2 (Supplemental Material) show Cohen kappa and percent agreement respectively for trained and in-training human scorers as well as for U-Sleep algorithms, relative to the ‘gold’ scoring; values for 5, 3 and 2 sleep stage approaches are presented. As expected, the median U-Sleep algorithm and human-scorer agreement with ‘gold’ scoring varied depending on staging approach, with lower values for five-stage approach compared to three- and two-stage approaches. There was minimal difference in algorithm performance when using differing EEG leads.

**Table 1:** Median (inter-quartile range) sleep staging agreement (assessed by Cohen’s kappa) of U-Sleep algorithms and human scorers with the gold standard for the concordance dataset of 50 PSG excerpts.

|  | **Five stage**<br>**approach** | **Three stage**<br>**approach** | **Two stage**<br>**approach\*** |
| --- | --- | --- | --- |
| *Trained Humans* | 0.78 (0.13) | 0.82 (0.11) | 0.86 (0.16) |
| *In Training Humans* | 0.73 (0.22) | 0.75 (0.17) | 0.78 (0.18) |
| *Pooled (all) Humans* | 0.77 (0.14) | 0.79 (0.10) | 0.82 (0.16) |
| *U-Sleep (F4)* | 0.76 (0.18) | 0.81 (0.34) | 0.87 (0.19) |
| *U-Sleep (C4)* | 0.78 (0.17) | 0.83 (0.19) | 0.89 (0.16) |
| *U-Sleep (O2)* | 0.76 (0.19) | 0.79 (0.26) | 0.89 (0.19) |
| *U-Sleep 2.0 (F4)* | 0.75 (0.20) | 0.77 (0.21) | 0.76 (0.19) |
| *U-Sleep 2.0 (C4)* | 0.77 (0.20) | 0.79 (0.25) | 0.77 (0.25) |
| *U-Sleep 2.0 (O2)* | 0.79 (0.19) | 0.79 (0.22) | 0.75 (0.26) |
| *U-Sleep 2.0 EEG (F4)* | 0.74 (0.16) | 0.75 (0.23) | 0.75 (0.25) |
| *U-Sleep 2.0 EEG (C4)* | 0.76 (0.17) | 0.76 (0.25) | 0.74 (0.24) |
| *U-Sleep 2.0 EEG (O2)* | 0.74 (0.23) | 0.79 (0.25) | 0.74 (0.42) |

*Note: Numbers presented are median (interquartile range), as the underlying distribution of Cohen's kappa values represented by each number does not appear to be normally distributed according to Shapiro Wilk test*
*\* 29/50 excerpts were excluded from the two-stage approach analysis as they had <5% wake epochs*

Wilcoxon TOST to assess for equivalence between trained scorer and U-Sleep algorithm performance determined that all tested U-Sleep algorithm variants (regardless of EEG lead used) for five sleep staging performed equivalently to trained human scorers, with the equivalence bounds set as within trained scorer IQR.

U-Sleep 2.0 and human scorer agreement with the ‘gold’ is depicted for each of the 50 excerpts in Figure 1. This figure shows that whilst there is congruence between trained and in-training humans and the U-Sleep algorithm in the majority of instances, there are multiple cases where in-training humans perform distinctly lower than trained humans (i.e. 2, 7,8, 16 and 28), and there are multiple instances where U-Sleep 2.0 algorithms perform distinctly worse than human scorers (i.e. 4, 44 and 48). Histograms displaying the number of PSG excerpts that fall into Cohen’s kappa agreement ranges when scored by U-Sleep 2.0 (using occipital EEG leads) and trained human scorers are shown in Figure 2, with additional histograms in Figure S1 (Supplemental Material) displaying algorithmic performance using other EEG leads and for humans subdivided into ‘trained’ and ‘in-training’ groups. Confusion matrices showing correct and incorrect classification of sleep stages by human scorers and U-Sleep algorithms are also shown in the Supplemental Material, in Figure S2.

**Figure 1:**
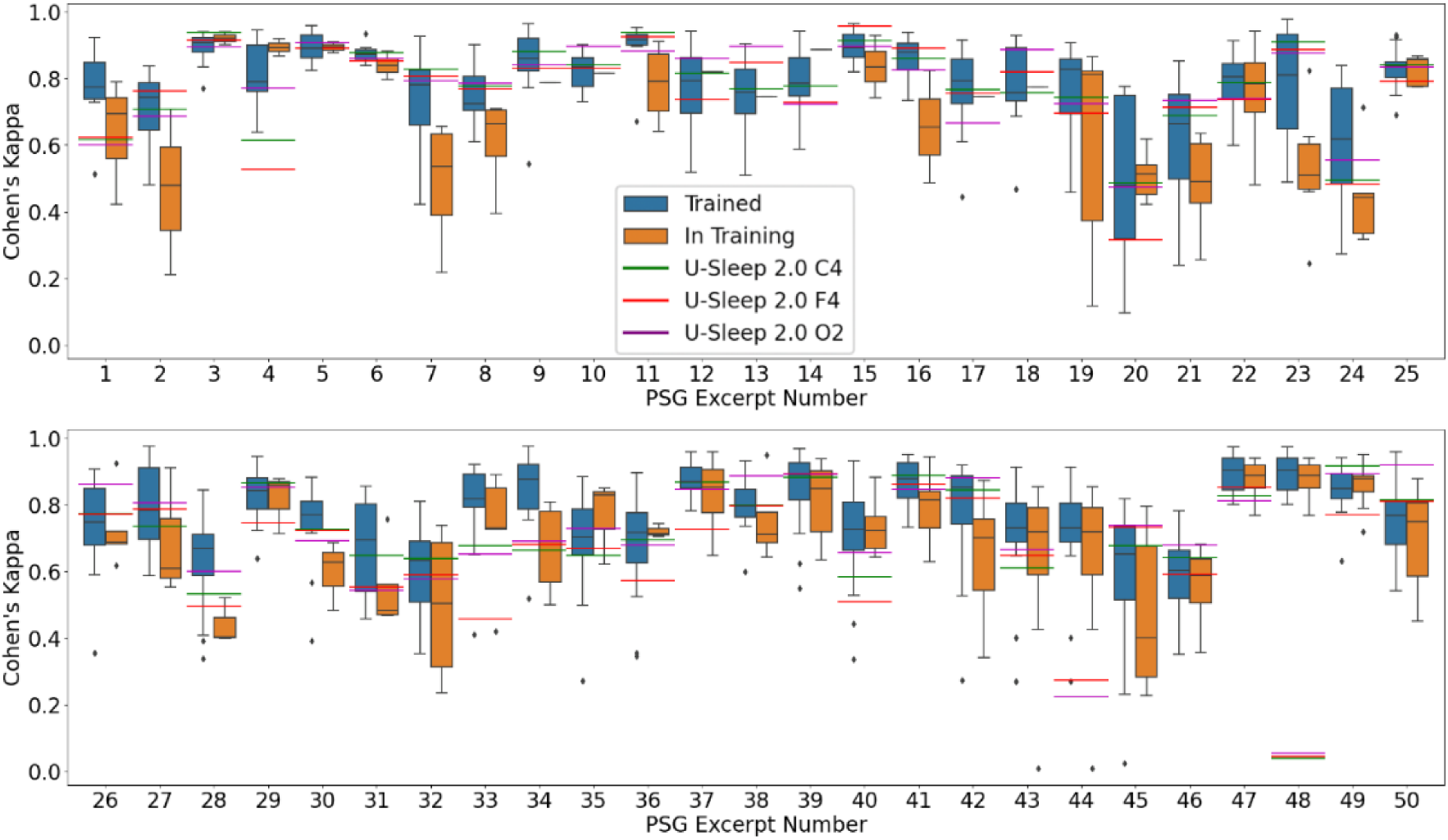
U-Sleep 2.0 and human scorer performance for five-stage pediatric sleep staging across PSG excerpts from concordance dataset.

**Figure 2:**
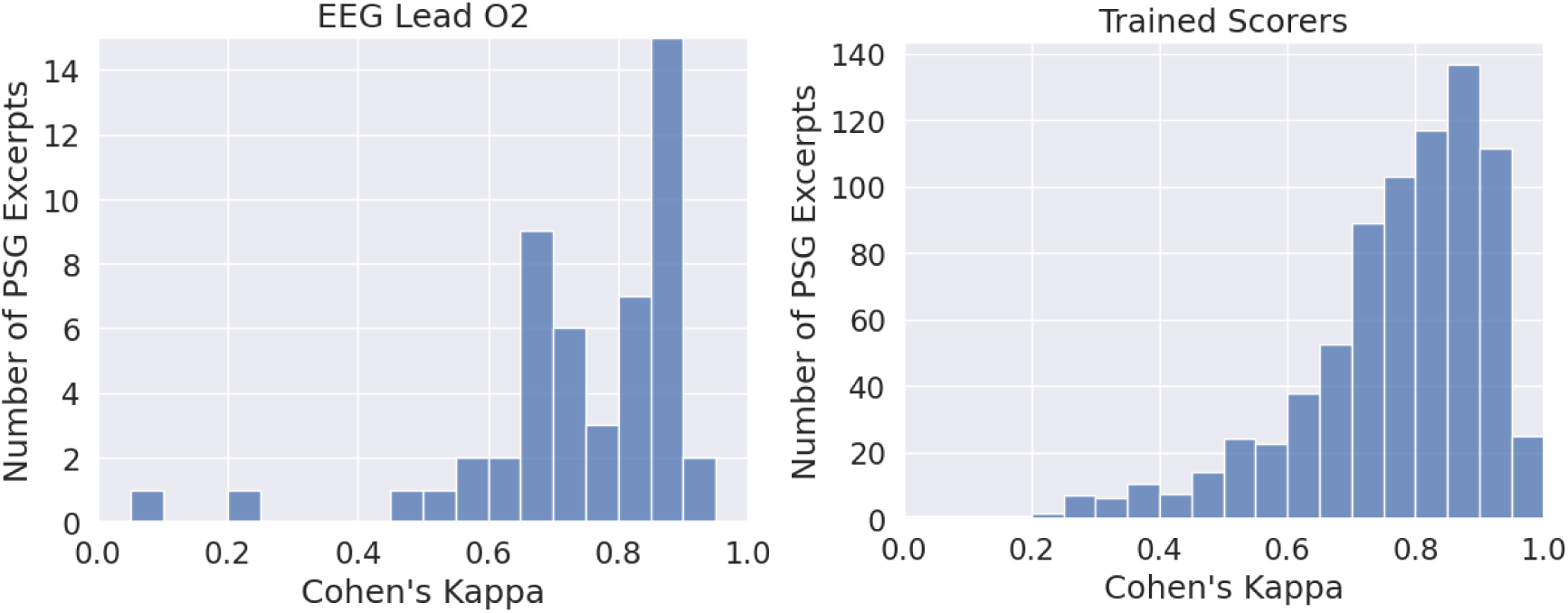
Histograms showing number of PSG excerpts with particular Cohen’s kappa values obtained by U-Sleep 2.0 (left) and trained scorers with weighting* (right) for five-stage sleep staging *\* As a variable number of human scorers partook in the scoring of each PSG excerpt, the weighting system compensates for this by proportionally increasing the weighting of a human scoring a PSG excerpt with less human scorers compared to another with more, in order for each of the 50 PSG excerpts to contribute equally to the histogram.* *Note: gold standard was defined as trained expert majority score*.

### Clinical Dataset

Analysis of U-Sleep 2.0 agreement with the ‘gold’ scoring from the clinical dataset showed median Cohen’s kappa varied slightly with sleep stage approach, and percent simple agreement increased when moving from five stage to three and two stage approaches (Table 2 & Table S2 in Supplemental Material). We found age is likely to have a substantial, non-linear impact on the performance of the U-Sleep model, particularly in infancy and early childhood (Figure 3). Generalised additive modelling of U-Sleep performance that included adjustment for this effect of age showed sleep efficiency, primary diagnosis category (Figure 4) and significant SDB (Figure 5) impacted further on U-Sleep performance in a linear fashion. Multi-variable linear modelling suggests each of these effects are independent of the other (Table 3). Notably, when also adjusted by age, central, moderate-severe obstructive or mixed SDB is associated with a 0.10-0.11 reduction in Kappa agreement; and the presence of a disorder known to likely impact EEG morphology is associated with a 0.15 reduction in Kappa agreement.

**Figure 3:**
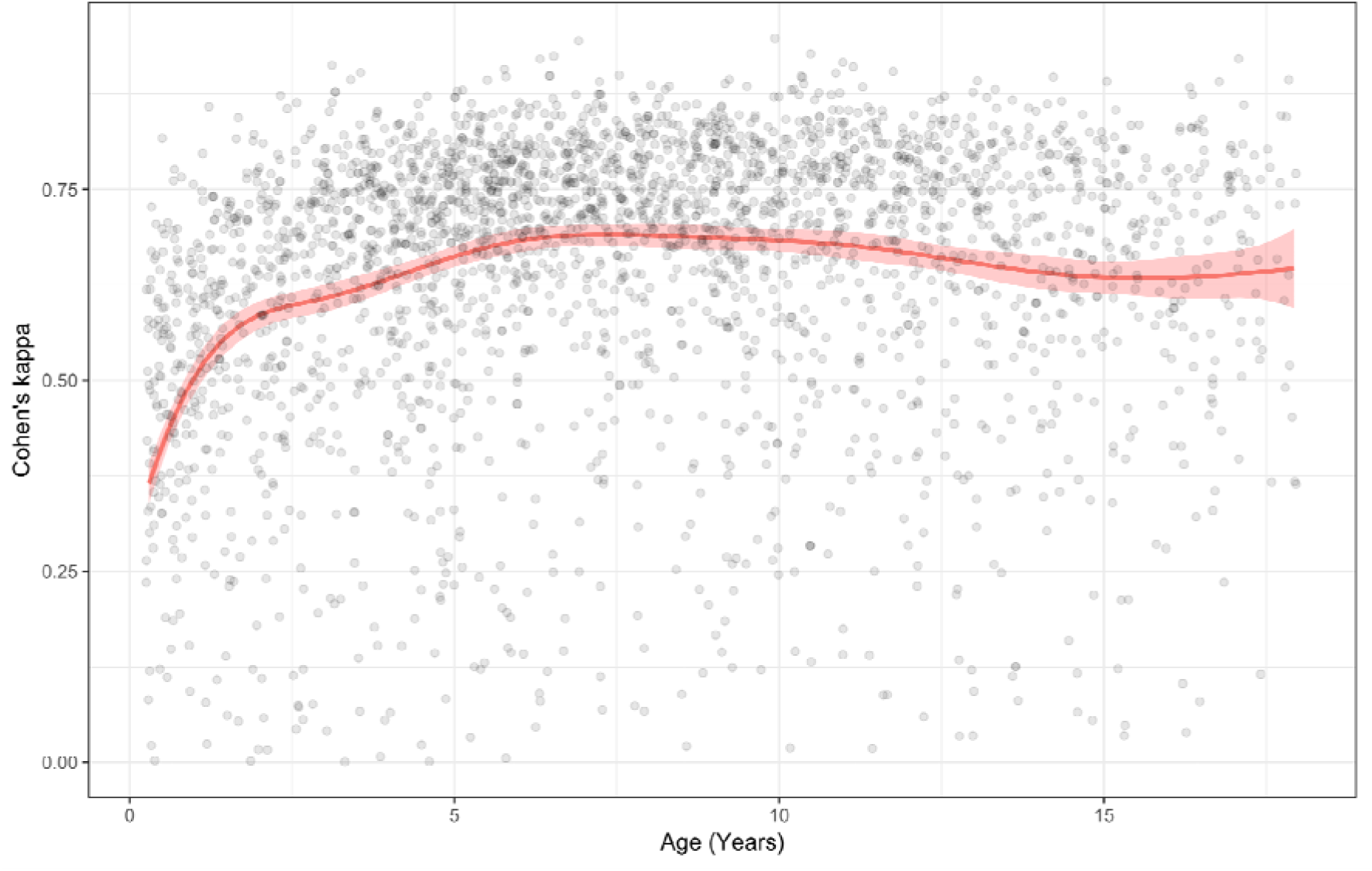
Generalized additive modelling of Cohen’s kappa using a P-spline of age (3 months-18 years).

**Figure 4:**
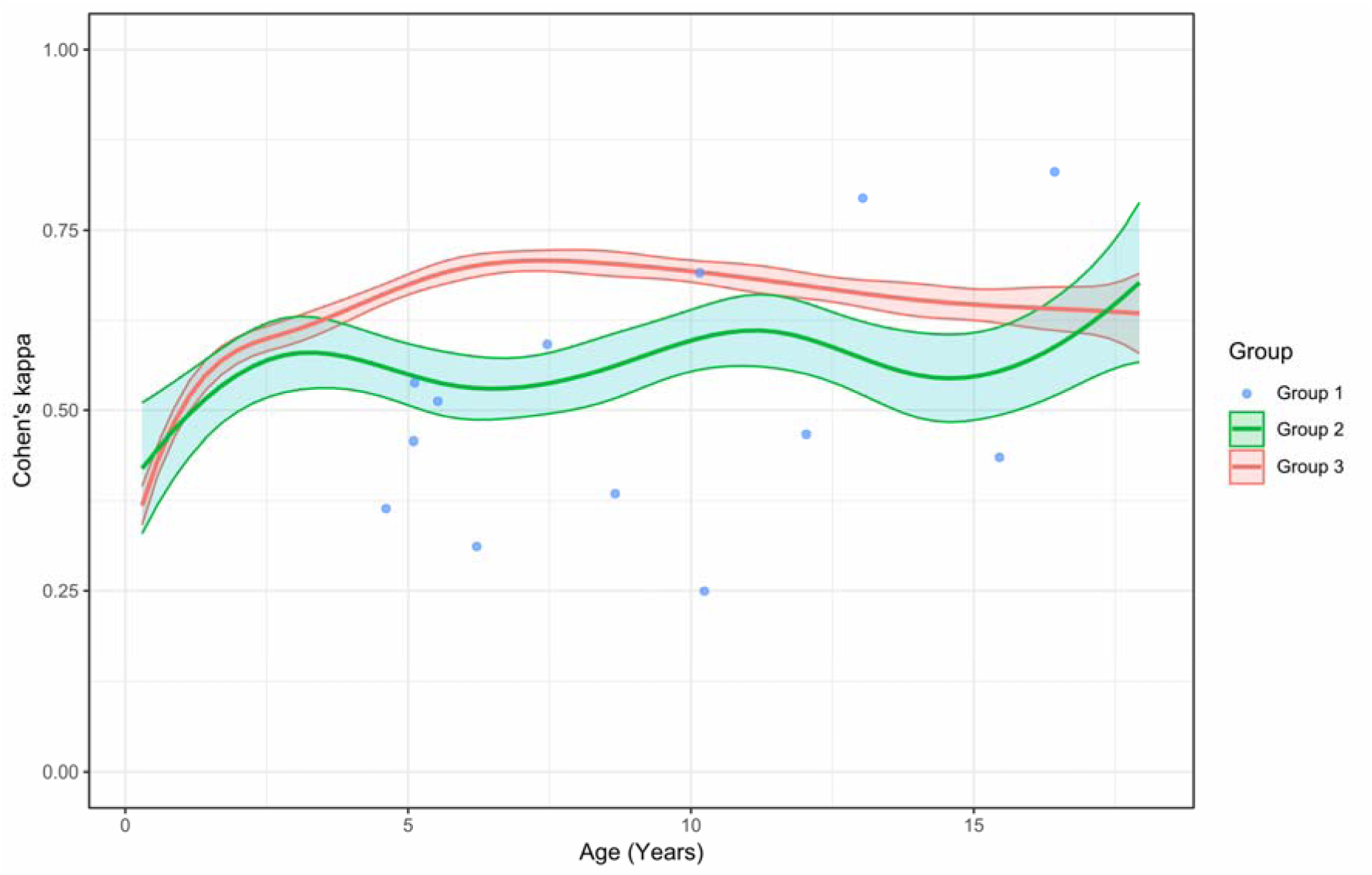
Generalized additive modelling of Cohen’s kappa using a P-spline of age for children grouped by comorbid diagnosis category *Group 1: comorbid diagnosis highly likely to manifest with sleep EEG abnormalities* *Group 2: comorbid diagnosis that may cause sleep EEG abnormalities* *Group 3: remaining children*

**Figure 5:**
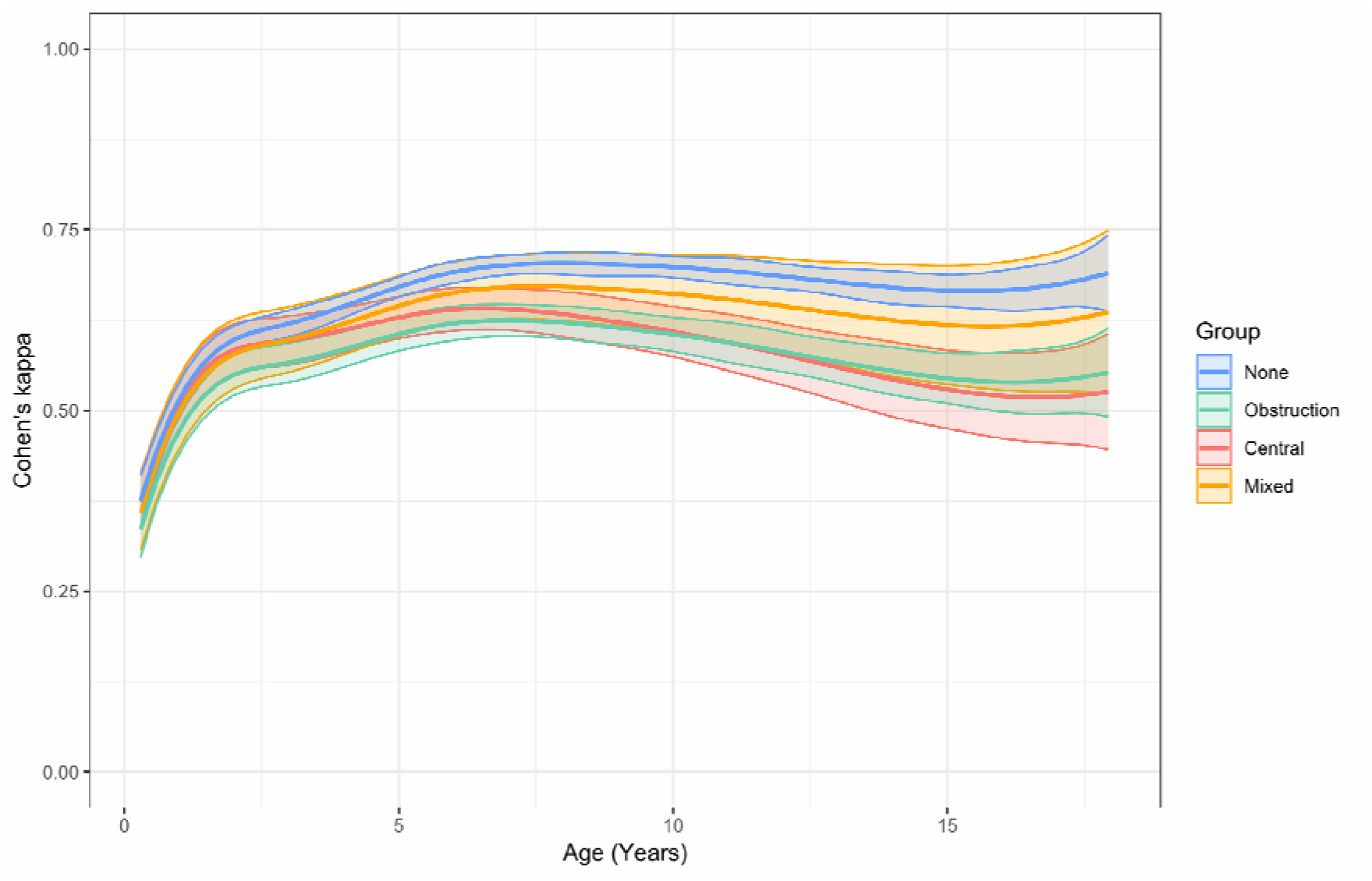
Generalized additive modelling of Cohen’s kappa using a P-spline of age for children with central, moderate-severe obstructive, or mixed sleep disordered breathing.

**Table 2:**
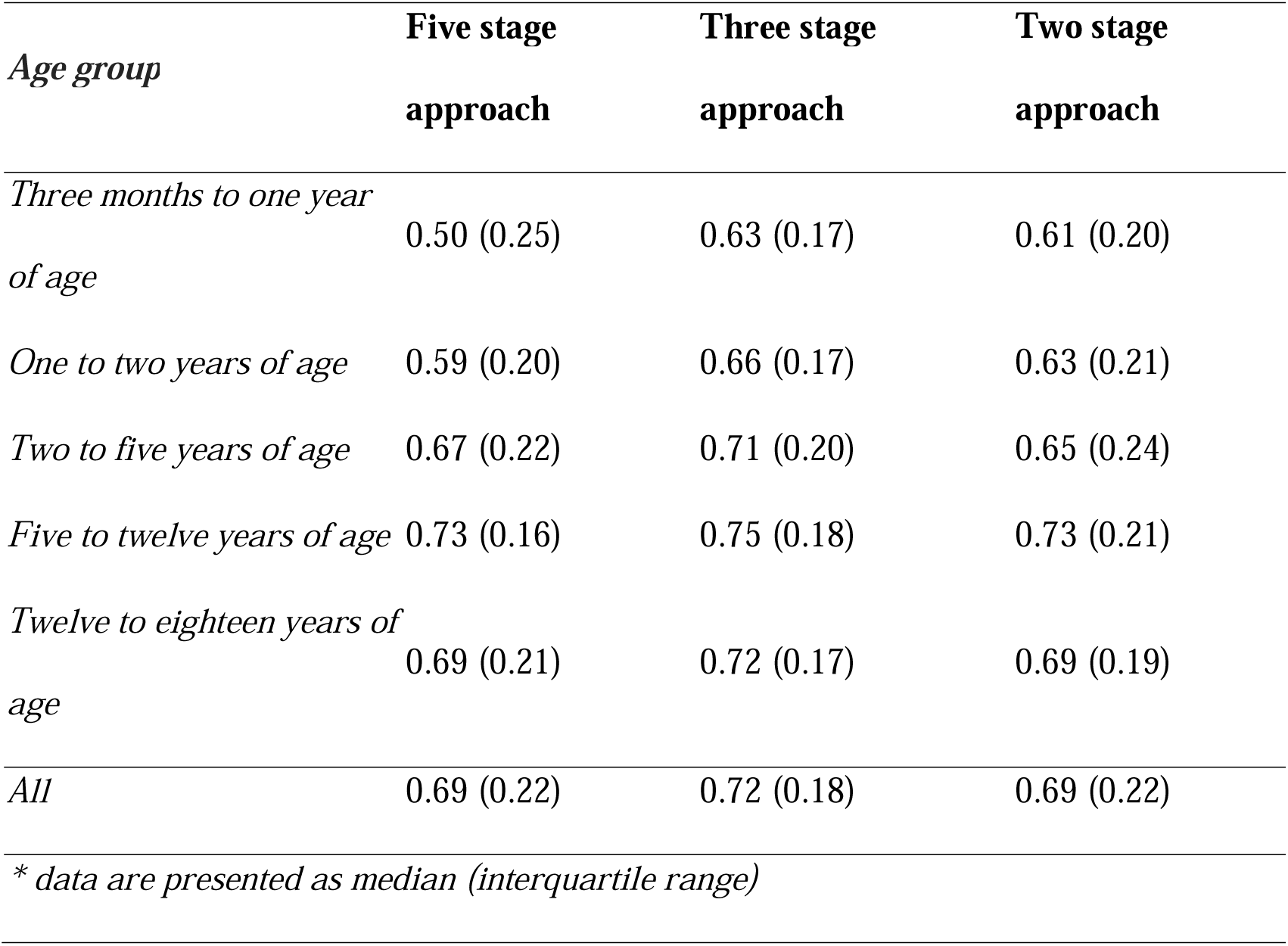
Sleep staging agreement (assessed by Cohen’s kappa) of U-Sleep 2.0 with the ‘gold standard’ epoch labels for the clinical dataset by patient age group.

| *Age group* | **Five stage**<br>**approach** | **Three stage**<br>**approach** | **Two stage**<br>**approach** |
| --- | --- | --- | --- |
| *Three months to one year*<br>*of age* | 0.50 (0.25) | 0.63 (0.17) | 0.61 (0.20) |
| *One to two years of age* | 0.59 (0.20) | 0.66 (0.17) | 0.63 (0.21) |
| *Two to five years of age* | 0.67 (0.22) | 0.71 (0.20) | 0.65 (0.24) |
| *Five to twelve years of age* | 0.73 (0.16) | 0.75 (0.18) | 0.73 (0.21) |
| *Twelve to eighteen years of*<br>*age* | 0.69 (0.21) | 0.72 (0.17) | 0.69 (0.19) |
| *All* | 0.69 (0.22) | 0.72 (0.18) | 0.69 (0.22) |
| *\* data are presented as median (interquartile range)* |  |  |  |

**Table 3:**
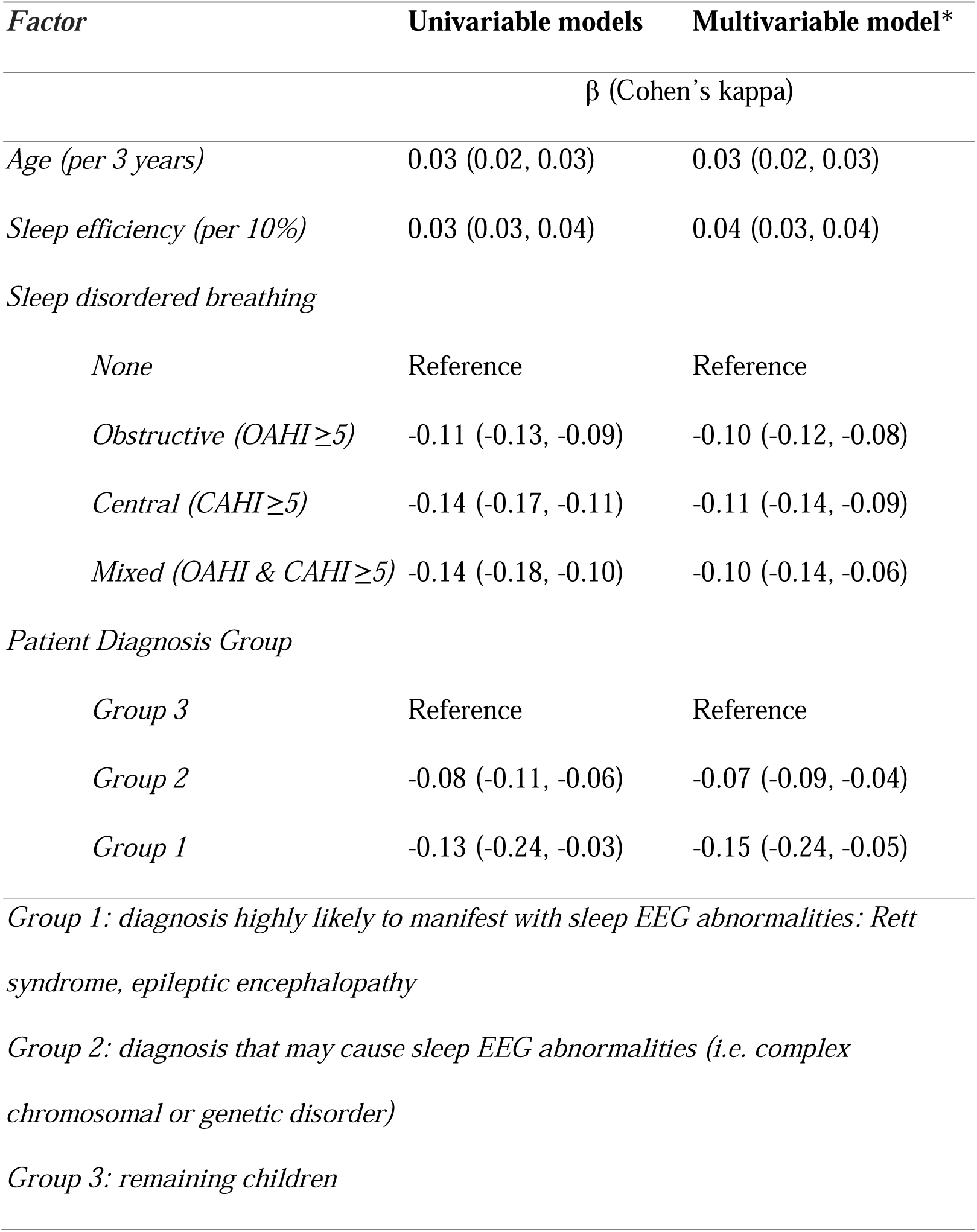
Univariable and multivariable modelling of factors affecting U-Sleep 2.0 5-stage performance.

| *Factor* | **Univariable models** | **Multivariable model\*** |
| --- | --- | --- |
| | $\beta$ (Cohen's kappa) | |
| *Age (per 3 years)* | 0.03 (0.02, 0.03) | 0.03 (0.02, 0.03) |
| *Sleep efficiency (per 10%)* | 0.03 (0.03, 0.04) | 0.04 (0.03, 0.04) |
| *Sleep disordered breathing* |  |  |
| *None* | Reference | Reference |
| *Obstructive (OAHl $\geq 5$)* | -0.11 (-0.13, -0.09) | -0.10 (-0.12, -0.08) |
| *Central (CAHI $\geq 5$)* | -0.14 (-0.17, -0.11) | -0.11 (-0.14, -0.09) |
| *Mixed (OAHl & CAHI $\geq 5$)* | -0.14 (-0.18, -0.10) | -0.10 (-0.14, -0.06) |
| *Patient Diagnosis Group* |  |  |
| *Group 3* | Reference | Reference |
| *Group 2* | -0.08 (-0.11, -0.06) | -0.07 (-0.09, -0.04) |
| *Group 1* | -0.13 (-0.24, -0.03) | -0.15 (-0.24, -0.05) |
| *Group 1: diagnosis highly likely to manifest with sleep EEG abnormalities: Rett syndrome, epileptic encephalopathy* |  |  |
| *Group 2: diagnosis that may cause sleep EEG abnormalities (i.e. complex chromosomal or genetic disorder)* |  |  |
| *Group 3: remaining children* |  |  |

## Discussion

Automated pediatric sleep staging tools have been described^17–25^ and are potentially clinically useful, but to date have not been independently tested or integrated into clinical care. The aim of this study was to independently test publicly available versions of the U-Sleep sleep staging algorithm and compare the performance of these models to clinician expert scoring in pediatric data. Our objectives were to: (1) compare the performance of U-Sleep algorithms to human inter-scorer performance relative to a trained expert majority-score gold standard; and (2) to identify if there are key clinical or demographic characteristics which impact the accuracy of the U-Sleep algorithm. Using 50 pediatric PSG excerpts, U-Sleep algorithm performance was found by statistical equivalence testing to be similar to that of trained scorers, although some outliers were noted. In a large retrospective clinical dataset, we identified that the agreement between clinician sleep staging and a chosen algorithm U-Sleep 2.0 was adversely impacted by age (particularly age less than two years), the presence of significant SDB, and clinical disorders that likely impact EEG morphology (with reductions in kappa of ≥0.1). In summary, independent evaluation indicates that U-Sleep performance is likely to be adequate for pediatric clinical application for most cases provided there is appropriate trained human review. Caution should be applied when considering use for younger children, those with significant SDB, and those with medical conditions likely to cause EEG abnormality.

Our evaluation of the performance of a freely available pediatric sleep staging solution, U-Sleep, found that algorithm concordance was similar to that of trained human scorers when tested against 50 different PSG excerpts, where the ‘gold’ was derived from scoring performed by multiple expert pediatric sleep clinicians from different pediatric sleep centers. However, in three (6%) instances, U-Sleep algorithm performance, even when using a different EEG lead input, was consistently a negative outlier i.e. had notably poorer performance than trained humans. The ‘black box’ nature of neural networks such as those used in U-Sleep makes it difficult to determine why errors in classification are made by the algorithm. Whilst explainable AI approaches for pediatric sleep staging designed to provide greater insight into how an algorithm performs its classification tasks have been described,^18^ these are not freely available. We therefore explored how demographic, clinical and PSG features were associated with poorer U-Sleep performance, using a much larger clinical dataset from a single center (>3000 pediatric diagnostic PSGs). Whilst U-Sleep 2.0 algorithmic concordance with clinician epoch labelling for five-stage sleep scoring was substantial (median Cohen’s kappa = 0.69), age less than two years, diagnosis of Rett Syndrome or epileptic encephalopathy (conditions highly likely to cause abnormalities in sleep EEG), and the presence of ≥5 obstructive or ≥5 central events per hour were associated with poorer U-Sleep performance (each associated with decreases in kappa of ≥0.1).

There is biologic plausibility as to why U-Sleep algorithm performance is poorer in younger children compared to older ones. As infants and young children’s brains are undergoing rapid growth, myelination and expanding connectivity, it is well-recognized that their brainwave morphology differs to that of the older child and mature adult.^26^ Similarly, if a patient has frequent obstructive or central events (leading to cortical arousals observed in EEG) or a significant neurologic/genetic disorder, then EEG disruption and deviation of EEG from the baseline morphology can be expected, which in turn may impact U-Sleep algorithmic performance. This phenomenon may affect other sleep staging algorithms, and indeed a recent report of poorer automated sleep staging accuracy for children with drug-resistant epilepsy compared to children with well-controlled or no epilepsy using a different algorithm, SeqSleepNet, confirms this.^30^

Performance differences between U-Sleep versions were minor. There was a marginally lower median kappa (-0.02) in the 2.0 EEG-only model in comparison to the 2.0 model, with misclassification increase for wake epoch detection (Figure 3), suggesting that some value with regard to algorithmic performance may be attributable to the EOG signal. Unsurprisingly, N1 was the sleep stage with the poorest agreement, a phenomenon well-described in the existing literature,^31^ with markedly better U-Sleep agreement with the ‘gold’ for other stages.

A particular strength of our research is that the accuracy of U-Sleep algorithms for determining sleep stages in young children and in those with medical conditions that might impact their neural sleep architecture was specifically investigated. Undertaking such testing is advisable prior to any clinical implementation to minimize the risk of bias in health care.^32^ However, there are also limitations to our research. Our clinical dataset is limited to that performed at a single center. Agreement in sleep staging between expert scorers from different centers can vary, which means results may not be generalizable to sleep staging performed at all other sites.^33^ This limitation is mitigated by our analysis of the concordance dataset, in which the ‘gold’ is determined by a larger number of scorers who, by participating in a benchmarking exercise, are likely to be paying closer attention to sleep staging accuracy, which improves their performance.^34^ We speculate that this is the reason why U-Sleep performance is better in the context of the concordance dataset in comparison to the clinical dataset.

Addressing poorer automated sleep staging accuracy affecting particular pediatric subgroups is a challenge our research has identified. Whilst additional training data or customized models might improve algorithmic performance for some groups (e.g. those <2 years who may be poorly represented in existing training datasets), this approach may be less effective for increasing accuracy for others, such as those with significant SDB or underlying neurological conditions, because these conditions are inherently linked to EEG signal disruption/abnormality. Indeed, increasing frequency of apnea, hypopnea and arousal events has been shown to adversely affect inter-clinician concordance,^33^ not just algorithmic sleep staging concordance. Therefore, if algorithmic pediatric sleep staging was to be adopted into clinical use, we would recommend a) review of all automated staging by a trained human expert, and b) particular caution be applied to use for children <2 years old, with frequent obstructive and/or central events, or with medical conditions that predispose to abnormal sleep EEG.

For most children, the accuracy of automated sleep staging using U-Sleep algorithms is comparable to human scorers. Barriers to adoption in pediatric sleep medicine can now be considered less attributable to poor algorithmic performance alone and include implementation hurdles such as licensing requirements, challenges integrating with existing software, pediatric clinician perception of the acceptability of artificial intelligence-backed algorithms being used in their field, and ensuring use that takes into account variability in pediatric patients. For specific groups, such as infants/young children and children with conditions affecting brainwave morphology, algorithm performance is poorer. Therefore, development of tailored algorithms may be justified for specific groups, and at this stage expert clinician review of automated pediatric sleep staging is recommended if it is to be used clinically. Implementation of automated sleep staging for pediatric patients will need to consider the needs of end users i.e., clinicians and the patients in their care.^35^ Integration with commonly-used existing user interfaces alongside the development of adjustable, explainable models that suit a wide variety of children and allow for a degree of algorithm transparency and customization to aid a clinician decision-support approach is perhaps more likely to garner widespread acceptance and clinical uptake.

## Abbreviations

CAHI: central apnea hypopnea index
EOG: electrooculogram
EEG: electroencephalography
IQR: interquartile range
OLS: ordinary least squares
OAHI: obstructive apnea hypopnea index
PSG: polysomnography
REM: rapid eye movement
SDB: sleep disordered breathing
TOST: two one-sided test

## Data Availability

Data produced in the present study are available upon reasonable request to the authors

## Supplemental Material

**Figure S1:**
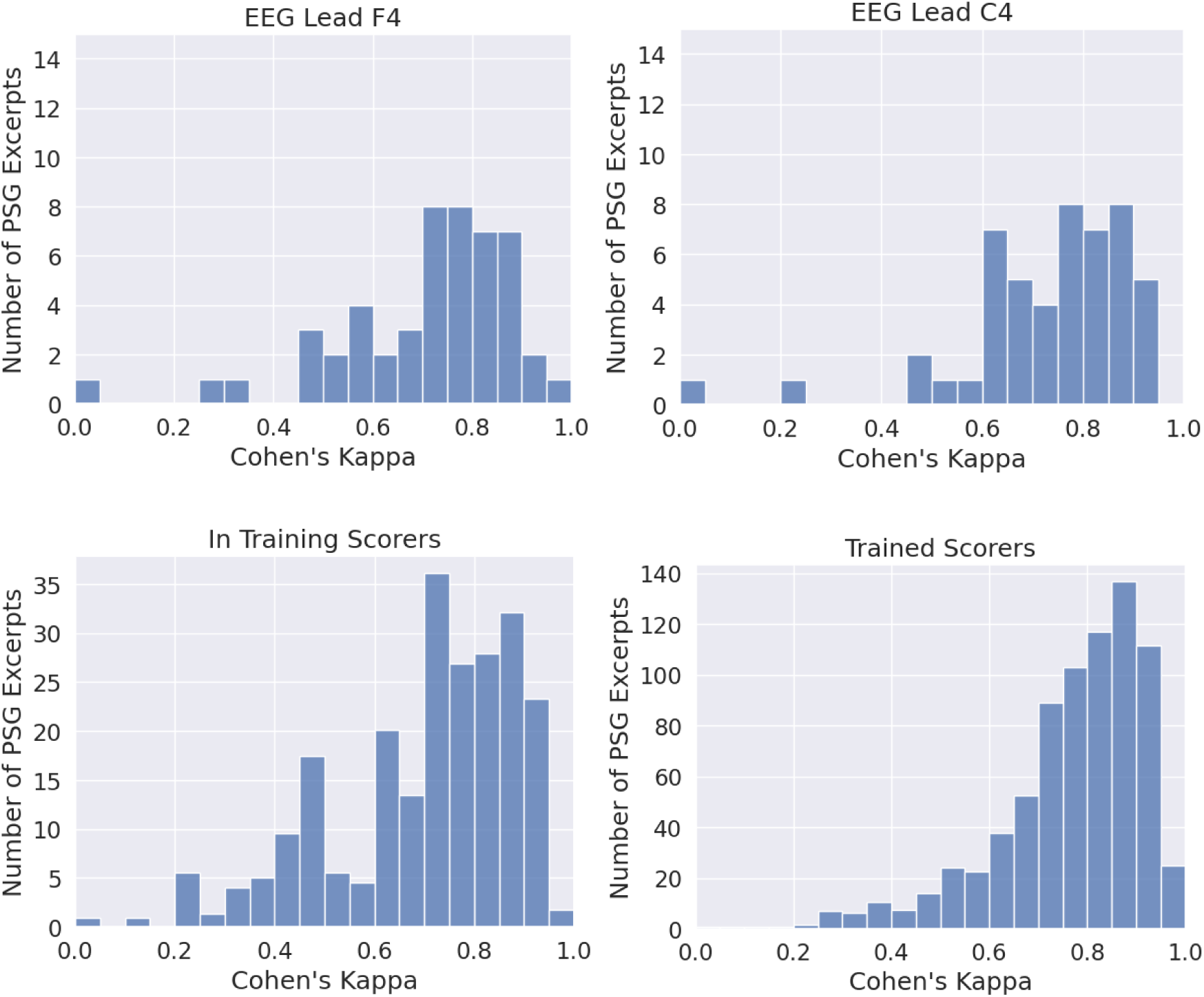
Histograms showing number of PSG excerpts from the concordance dataset with particular Cohen’s kappa values obtained, when scored by U-sleep 2.0 on different EEG leads (top), and in-training and trained human scorers with a weighting applied* (bottom) for the task of five-stage sleep staging; gold standard defined as trained expert majority-score. *\* As a variable number of human scorers partook in the scoring of each PSG excerpt, the weighting system compensates for this by proportionally increasing the weighting of a human scoring a PSG excerpt with less human scorers compared to another with more, in order for each of the 50 PSG excerpts to contribute equally to the histogram*.

**Figure S2:**
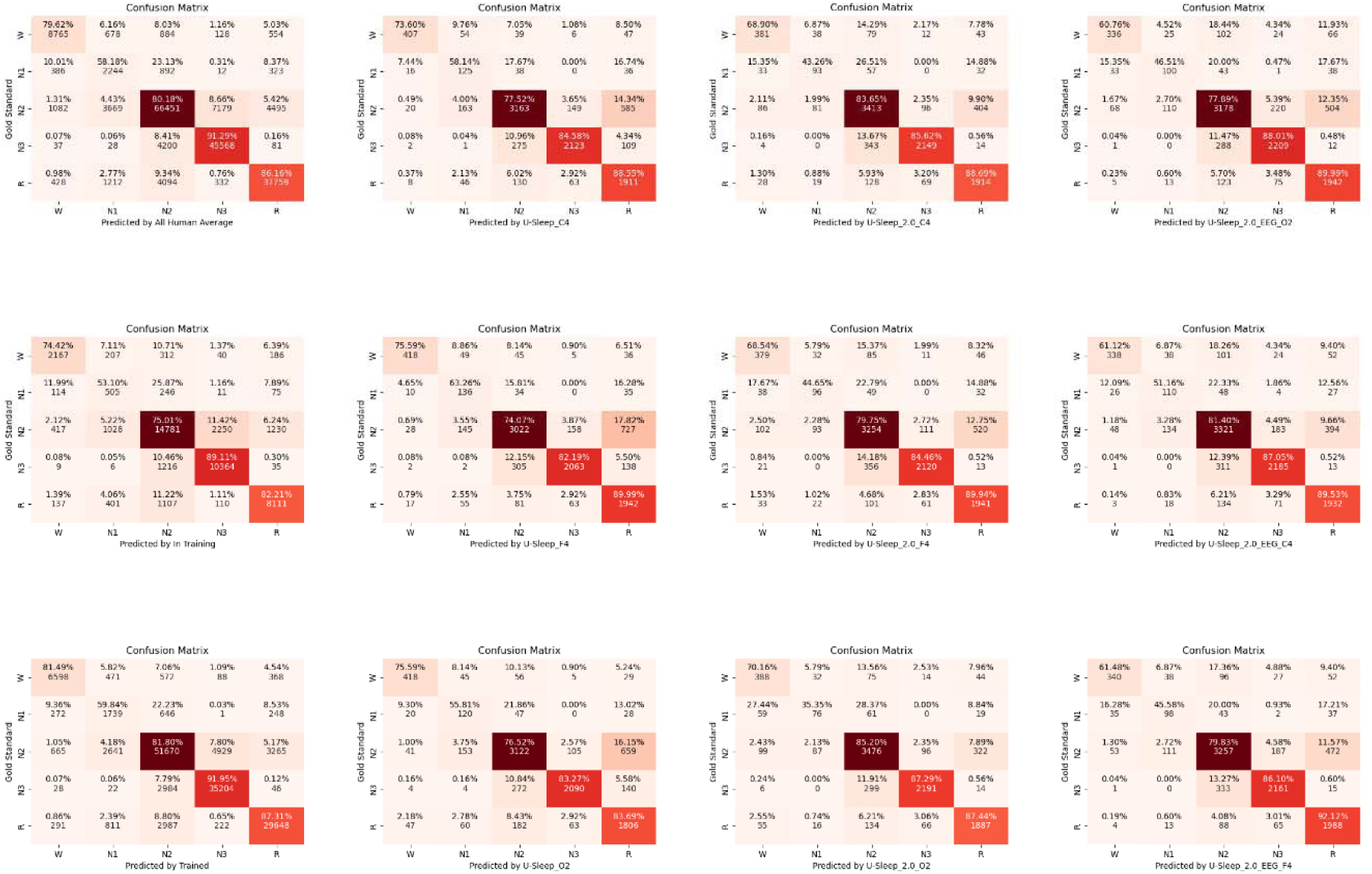
Confusion matrices showing correct and incorrect classification of sleep stages by U-Sleep algorithms and human scorers

**Table S1:** Number and type of epochs available in the clinical dataset by patient age.

| **Age** | **REM** | **N1** | **N2** | **N3** | **Wake** |
| --- | --- | --- | --- | --- | --- |
| **3m - 1y** | 39787 | 3572 | 85953 | 45410 | 34440 |
| **1-2y** | 42530 | 3642 | 76072 | 48244 | 39039 |
| **2-5y** | 150849 | 9701 | 290417 | 154732 | 136047 |
| **5-12y** | 330327 | 21587 | 626213 | 347144 | 39039 |
| **12-18y** | 148342 | 8851 | 272557 | 153253 | 124341 |
| **Total** | 729256 | 48214 | 1371568 | 763010 | 387511 |

**Table S2:** Median (inter-quartile range) sleep staging concordance (assessed by simple agreement) of A) U-Sleep algorithms and human scorers with the gold standard for the concordance dataset of 50 PSG excerpts and B) U-Sleep 2.0 with the ‘gold standard’ epoch labels for the clinical dataset by patient age group

|  | **Five stage**<br>**approach** | **Three stage**<br>**approach** | **Two stage**<br>**approach\*** |
| --- | --- | --- | --- |
| **A. Concordance Dataset** |  |  |  |
| *Trained Human Average* | 0.87 (0.08) | 0.94 (0.05) | 0.98 (0.02) |
| *In Training Human Average* | 0.83 (0.14) | 0.92 (0.08) | 0.97 (0.04) |
| *Pooled (all) Human Average* | 0.85 (0.08) | 0.94 (0.05) | 0.97 (0.02) |
| *U-Sleep (F4)* | 0.86 (0.09) | 0.92 (0.09) | 0.98 (0.03) |
| *U-Sleep (C4)* | 0.87 (0.09) | 0.93 (0.10) | 0.98 (0.03) |
| *U-Sleep (O2)* | 0.86 (0.12) | 0.92 (0.11) | 0.97 (0.04) |
| *U-Sleep 2.0 (F4)* | 0.86 (0.11) | 0.91 (0.08) | 0.96 (0.05) |
| *U-Sleep 2.0 (C4)* | 0.86 (0.13) | 0.93 (0.10) | 0.96 (0.04) |
| *U-Sleep 2.0 (O2)* | 0.88 (0.11) | 0.93 (0.07) | 0.96 (0.04) |
| *U-Sleep 2.0 EEG (F4)* | 0.84 (0.09) | 0.92 (0.11) | 0.96 (0.05) |
| *U-Sleep 2.0 EEG (C4)* | 0.85 (0.10) | 0.93 (0.11) | 0.96 (0.05) |
| *U-Sleep 2.0 EEG (O2)* | 0.86 (0.15) | 0.92 (0.13) | 0.97 (0.06) |
| **BA. Clinical Dataset** |  |  |  |
| *Age three months to one year* | 0.69 (0.15) | 0.78 (0.11) | 0.91 (0.07) |
| *Age one to two years* | 0.73 (0.12) | 0.82 (0.09) | 0.92 (0.07) |
| *Age two to five years* | 0.77 (0.13) | 0.85 (0.10) | 0.93 (0.06) |
| *Age five to twelve years* | 0.80 (0.10) | 0.87 (0.07) | 0.94 (0.05) |
| *Age twelve to eighteen years* | 0.78 (0.12) | 0.86 (0.09) | 0.93 (0.07) |
| *Total (all ages pooled)* | 0.78 (0.13) | 0.86 (0.10) | 0.93 (0.06) |
| *Note: Numbers presented are median (interquartile range), as the underlying distribution of simple agreement values represented by each number did not appear to be normally distributed according to Shapiro Wilk test*<br>*\* 29/50 excerpts from concordance dataset only were excluded from two-stage approach analysis as they had <5% wake epochs* |  |  |  |

